# Maternal risk factors contributing to stillbirths in semi-rural Ghana: Insights from a cross-sectional study

**DOI:** 10.1101/2023.10.25.23297496

**Authors:** Silas Adjei-Gyamfi, Abigail Asirifi, Gafaru Mamudu, Wisdom Peprah, Clotilda Asobuno, Constance Siakwan Dapaah, Paul Armah Aryee

## Abstract

**Background:** Stillbirth remains a critical public health challenge in low- and middle-income countries, impeding progress toward achieving the Sustainable Development Goals. Given the diverse dynamics of the prevalence and associated factors of stillbirths in various low-resource settings, such as Ghana, it is essential to have comprehensive knowledge to address and mitigate these factors that could influence stillbirths. However, there is a paucity of data on maternal factors contributing to stillbirths in semi-rural areas of Ghana.

**Objective:** This study aimed to determine the prevalence and associated maternal factors of stillbirths in semi-rural Ghana.

**Design:** This is a cross-sectional study that utilised a quantitative approach.

**Methods:** The study was conducted among 422 mothers who delivered in public health facilities and reside in the Savelugu Municipality of Northern Ghana. Sociodemographic, obstetric, and medical-related data were gathered through structured interviews and from record reviews. Chi-square test of proportions and multiple logistic regression analyses were employed to identify the independent factors associated with stillbirths at a 95% confidence level.

**Results:** Stillbirth rate was estimated at 4.0% (95%CI: 2.4%–6.4%). The analysis revealed that anaemia in the third trimester of pregnancy (aOR:3.15; 95%CI:2.94–6.10; p=0.034), positive sickle cell status (aOR:2.91; 95%CI:2.50–5.01; p=0.018), and non-utilisation of insecticide-treated bed nets (ITNs) (aOR:7.75; 95%CI:4.33–8.80; p=0.001) during pregnancy were significantly associated with increased risk of stillbirths.

**Conclusion:** The relatively high prevalence of stillbirths in the municipality was influenced by medical factors (particularly anaemia and positive sickle cell status) and unsafe health practices via the non-use of ITNs during pregnancy. The management of the Ghana Health Service, alongside program planners and other stakeholders involved in enhancing perinatal and maternal health, should focus on implementing effective initiatives such as improved health education on nutrition and safety practices at the facility and community levels, as well as robust referral systems to prevent and reduce stillbirths in semi-rural Ghana.

**Plain language summary:** Stillbirth, which is the death of a baby occuring before or during childbirth, not only remain a public health issue but also cause emotional distress for mothers in most African countries. Despite the importance of understanding the maternal factors of stillbirths, there is limited data available, especially in semi-rural areas of Ghana. A study was therefore undertaken to assess the prevalence and maternal factors associated with stillbirths in semi-rural Ghana. This study involved women who gave birth in public health facilities and live in the Savelugu Municipal area. Researchers collected data through structured interviews and from medical records. The data were analysed using chi-square tests and logistic regression to identify maternal factors linked to stillbirths. The findings showed a stillbirth rate of 4.0%. Key factors associated with stillbirths included maternal anaemia in the third trimester of pregnancy, positive sickle cell status, and not using insecticide-treated bed nets (ITNs) during pregnancy. The study highlights the need for improved health education on nutrition and safe health practices to reduce and/or prevent stillbirths. Priority should be given to managing medical conditions such as anaemia in pregnancy and sickle cell disease, alongside promoting the use of ITNs to safeguard pregnant women against malaria and subsequently reduce the risk of stillbirths.

## Introduction

Stillbirth remains a critical public health challenge in low- and middle-income countries (LMICs), impeding progress toward achieving the Sustainable Development Goals (1–3). Defined by the World Health Organisation (WHO), stillbirth is denoted as the birth of a baby with no signs of life occurring at 28 weeks or more of gestation (3), and it accounts for more than half of all perinatal and maternal mortalities worldwide (4). Stillbirths, also called stillborns, trigger maternal psychological trauma which could affect the socio-economic growth and development in LMICs (1–3).

The aetiology and antecedents of stillbirths are multifactorial and diverse, influenced by sociological, physiological (medical), and physical factors (1). It is reported that the stillbirth rate and its determinants are dependent on the geographic and sociodemographic distribution of the population (3,5). Therefore, the study findings from one setting might not apply to another. Universally, earlier studies acknowledged that the factors associated with stillbirths included congenital foetal deformities, foetal distress, preterm births, breech delivery, abruptio placentae, and male foetus (6–9). In most LMICs, pregnancy-related smoking and alcohol intake, maternal low socioeconomic status, multiparity, non-use of insecticide-treated bed nets (ITNs) for malaria prevention during pregnancy, maternal age less than 20 years, unmarried mothers, employment status, and delayed vaginal delivery were also regarded as some determinants of stillbirths in other past studies (9–13). In other low-resource settings, inadequate knowledge about antenatal care (ANC) services, cost, distance and the time needed to access care are major barriers to the effective uptake of quality ANC and particularly intrapartum services, which contributes significantly to stillbirths (14,15).

On a global scale, an estimated 2·6 million babies were born with no signs of life in 2015, which reduced to about two million stillborns (13.9 per 1000 births) in 2019 (5,16). LMICs particularly, sub-Saharan Africa (SSA) has the highest stillbirth rate globally, estimated at 21.7 per 1000 births, compared to 3.0 for high-income countries (HICs) (16) and accounts for more than one-third of the global burden of stillbirths (5,16). Furthermore, stillbirth rates vary widely across regions in the world, from 22·8 per 1000 births in Western and Central Africa to 2·9 in Western Europe. After West and Central Africa, Eastern and Southern Africa and South Asia have the second (20.5 per 1000 births) and third-highest (18.2 per 1000 births) stillbirth rates (16). Estimates of stillbirths in China, South Africa, and Nigeria are found to be 9.0, 35.0 and 40.0 per 1000 births respectively (10,11,14,17). Hug et al. 2021 observed a 2·3% annual decline in the stillbirth rate globally, with a sluggish decline rate in most Asian and SSA countries (16). These findings may hinder SSA countries from reaching the Every Newborn Action Plan target of 12 per 1000 births by 2030.

The vast differences in stillbirth rates between HICs and LMICs have led to stillbirth being equally regarded as a global development metric. Similar to many other SSA nations, Ghana faces challenges in the routine and adequate monitoring and recording of stillbirths (5,18). Diverse surveys conducted in Ghana estimate the stillbirth rate to fluctuate between approximately 14 and 22 per 1000 births (18,19). Also, some population health surveillance and health facility data from different parts of the country indicate even higher rates. For instance, the Ghana Maternal Health Survey in 2017 observed that 68% of stillbirths occur in rural areas compared to urban areas (18). Other findings also revealed 34 stillbirths/1000 births in Navrongo of the Upper East Region, 59 stillbirths/1000 births in the Ashanti Region and 109 stillbirths/1000 births in a tertiary hospital in the Greater Accra Region (7,20,21), which makes these high stillbirth occurrences very worrisome in the country. While these longitudinal studies reported a wide range of health facility, neonatal, paternal and maternal factors, none have exclusively delved into the broad spectrum of maternal factors that contribute to stillbirths. Additionally, most of these longitudinal studies were conducted in rural and urban areas. Hence, a cross-sectional study to provide a snapshot of the current situation in semi-rural communities is non-existent. Again, to date, little is particularly investigated about maternal factors contributing to stillbirths in semi-rural Ghana.

On the other hand, epidemiological studies evaluating stillbirths frequently utilise the concept of perinatal deaths, which encompasses both stillbirths and early neonatal deaths (5,16). These studies often rely on population health surveillance, community-based oral post- mortem reports and intricate analytical modelling techniques. However, the utility of these estimates is constrained by the etiological distinctions between stillbirths and early neonatal deaths, particularly given that the intrapartum period represents the highest-risk phase for stillbirths. To achieve the World Health Assembly’s national target of a stillbirth prevalence of 12 or less per 1000 births by 2030 in every country through the Every Newborn Action Plan (3), the maternal factors of stillbirths and their dynamics in different settings or context must be well understood and appreciated. Although stillbirths are higher in LMICs, including Ghana (7,20), relevant information on the maternal factors of stillbirths is inadequately and incorrectly documented in the district and/or regional databank. Hence, adequate evidence of information on these factors in semi-rural Ghana could trigger useful interventions that may lead to preventable stillbirths and subsequently contribute to improving maternal and perinatal health.

The purpose of the study is to estimate the prevalence of stillbirths and examine the maternal factors associated with stillbirths in a semi-rural municipality in the Northern Region of Ghana. We examined the effect of maternal risk factors on stillbirths by utilising a broad spectrum of sociodemographic, medical or clinical and obstetric or antenatal characteristics of the mothers. Given the distinct and unique nature of this study, the findings will be crucial to inform clinical practice through timely interventions, decision-making, and policy ideation, with a particular emphasis on maternal factors in the semi-rural Ghana and SSA.

## Materials and methods

### Study design

This current survey is a cross-sectional study conducted among women who delivered in a semi-rural municipality in the Northern Region of Ghana. The design was employed to determine the prevalence and associated maternal factors of stillbirths. The study followed the STROBE checklist (22,23) of reporting (see Supplementary 1).

### Study setting

This study was conducted in Savelugu Municipality of Northern Ghana. Savelugu Municipality is one of the 16 districts/municipalities in the Northern Region of Ghana that shares boundaries with five neighbouring districts, including Gushegu, Karaga, Kumbungu, Nanton, and Tolon. The municipality has five major government (public) health facilities (one hospital and four health centres) that provide comprehensive maternal and newborn services, including free maternal and delivery services (24) to a population of approximately 140,000. In 2019, the coverage of ANC and postnatal care visits was 94% and 98%, respectively (25,26). Due to the increased average monthly delivery count of over 300 deliveries with higher records of unfavourable birth outcomes (25,27,28), this municipality was selected to investigate the factors of stillbirths among women who delivered in government health facilities.

### Sample size estimation

The sample size (n) was estimated using Cochran’s method with the WHO conservative design effect (DEF), 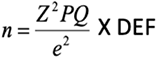 (29–31). A standard variate (Z) at a 95% confidence interval of 1.96, a standard error (e) of 5%, a prevalence (P) of stillbirths estimated at 21% in Northern Ghana (32), Q=1–P and WHO standard conservative DEF size of 1.5 (29,30), was applied to estimate the preliminary sample size at 384. An additional 10 percent was added to the calculated sample size to account for fall-outs, incomplete responses and damaged questionnaire (25,27,33) resulting in a final total sample of approximately 422.

### Study population

The study recruited a total of 422 women who visited the five major public facilities in the Savelugu Municipal for childbirth or delivery purposes from January 1 to June 31, 2020. All women who lived in the municipality, delivered at the municipal health facilities and had their health information recorded in the delivery registers were eligible for the study.

### Sampling

In the sampling stage, all five major government (public) health facilities were chosen for the study since all these facilities report on stillbirths every year (34). The sample sizes for each facility were determined using the probability proportional to size approach within a stratified random sampling framework (see Table 1). The delivery registers from each facility were used to line-list all eligible postpartum women (who delivered from January 1 to June 31, 2020) to form a sampling frame. A simple random sampling technique was then employed to select the required number of respondents from each facility. Specifically, a lottery method without replacement was utilised to ensure that each participant was selected only once.

**Table 1:** Description of proportionate sample size for study health facilities.

|  | Savelugu<br>Municipal<br>Hospital | Savelugu<br>Health<br>Centre | Pong Tamale<br>Health<br>Centre | Moglaa<br>Health<br>Centre | Diare<br>Health<br>Centre | Total |
| --- | --- | --- | --- | --- | --- | --- |
| Total half-year facility deliveries in 2020 (X) | 1,594 | 12 | 103 | 61 | 210 | 1,980 |
| Facility coverage (Y: X/1,980) | 80.5% | 0.6% | 5.2% | 3.1% | 10.6% | 100% |
| Approximate sample size (Z: Y x 422) | 339 | 3 | 22 | 13 | 45 | 422 |

The addresses and contact numbers of these selected respondents were recorded to trace them to their homes for the first part of data collection. Following home-based data collection, the information of recruited participants were re-traced to the delivery registers to complete the data collection at the designated health facilities. This approach was utilised to systematically reduce social desirability bias among the data collectors.

### Data collection

Data were collected using a pretested questionnaire by six well-trained data collectors (midwives and public health officers) from July 16 to September 31, 2020. The data collection was conducted during the period of COVID-19 pandemic, hence COVI-19 protocols were followed. The questionnaire was developed based on existing literature, previous studies, and sociological and biological plausibility (1,5,25,35). We verified the content validity of the questionnaire by consulting with experts in the field as well as the study authors (see Supplementary 2). Sociodemographic, obstetric or antenatal and medical-related information was collected through structured interviews and from record reviews (thus, delivery registers and maternal and child health record books [MCHRBs]). The data collectors were trained for one week on study purpose and background, sampling techniques, data collection procedures, ethical procedures and consent process among others. The data collection procedure was conducted in two phases. The first phase ran through a period of about two months and was conducted at the household level after sampling and contacting the eligible study participants. The second phase of the data collection lasted for about one month and was conducted at the health facilities level immediately after household-based data collection. This approach was relevant to reduce social desirability bias among the data collectors. It also mitigated selection bias among recruited participants by accounting for the non-attendance of health facilities by mothers who had experienced stillbirths.

Before the commencement of household data collection, the trained data collectors retrieved the addresses and contact information (telephone numbers) of eligible respondents and/or their representatives from the delivery registers with the support of the facility midwives. The data collectors arranged appointments and then visited the homes of the recruited respondents. Data such as age, marital status, education level, job status, ethnicity and religious affiliations were collected via structured interviews and were confirmed or validated in the MCHRBs. Information about first-trimester body mass index (since the foetal weight in the first trimester is negligible (36), antenatal visits, parity, gravidity, rhesus status, sickle cell status, tetanus-diphtheria (TD) immunisation status, haemoglobin levels across pregnancy trimesters, use of pre-pregnancy family planning, and utilisation of ITNs (1,28) were collected from their MCHRBs at their homes. Haemoglobin electrophoresis was not further conducted to confirm sickle cell variants by the health facilities. Following completion of the home-based data collection, information like pre-eclampsia diagnosis, postpartum haemorrhage, delivery mode, gestational age at birth and birth/delivery outcome prognosis (stillbirth or livebirth) based on the classification by health professionals (1,28) were immediately traced and extracted from the delivery registers at the health facilities level.

### Study variables

The outcome variable was birth outcome prognosis (stillbirths), categorised into binary outcomes for stillbirths (coded as 1) or livebirths (coded as 0), whereas exposure variables included all sociodemographic, obstetric or antenatal, and medical variables of the women, which were all categorised based on their normal distribution and existing studies (1,5,25,35).

According to the WHO, stillbirths were measured as the birth of a foetus at 28 or more weeks of gestation with no signs of life, while livebirths were regarded as newborns with signs of life (3). Haemoglobin levels were dichotomised into anaemia (< 11g/dl) and no anaemia (≥ 11g/dl). Gestational age at birth was grouped into < 37 weeks (preterm) and ≥ 37 weeks (normal-term). Parity was classified as primiparity (one delivery) and multiparity (two or more deliveries). Gravidity was grouped into primigravida (one pregnancy) and multigravida (two or more pregnancies) (26–28,33). Standardised WHO categories of pre-pregnancy body mass index (BMI) of underweight (<18.5 kg/m^2^ ), normal BMI (18.5–24.9 kg/m) and overweight (≥25.0 kg/m^2^) were utilised. Sickle cell and rhesus statuses were grouped as positive or negative, whereas ANC visits were also categorised into ≥ 8 visits and < 8 visits using the new 2016 WHO antenatal model which are also applicable to the Ghanaian health system (25,35).

### Statistical analysis

Frequencies and percentages were calculated for the sociodemographic, obstetric or antenatal and medical variables. Chi-square/Fisher’s analysis tests of proportions were employed to determine the statistical association between birth outcome prognosis (stillbirth) and all exposure variables at the bivariate level. These analyses were confirmed with univariate logistic regression. Multicollinearity among the significant exposure variables was then assessed with variance inflation factor (VIF), where variables with a VIF less than five were selected into multiple (binary) logistic regression analyses (25,26,28).

Applying simultaneous variable entry, the significant exposure variables with no multicollinearity issues were forwarded into the multiple logistic regression model and run to determine significant factors of stillbirths. All analyses were computed using STATA 17.0 (Stata Corporation, Texas, USA), and a p-value of less than 0.05 was considered significant during statistical analysis.

## Results

### Prevalence of stillbirths

The prevalence of stillbirths in the semi-rural municipality of Ghana is presented in Figure 1. All the data from the sampled 422 mothers were used for the analysis. The prevalence of stillbirth in this semi-rural municipality was estimated at 4.0% (95%CI: 2.4% – 6.4%).

**Figure 1.**
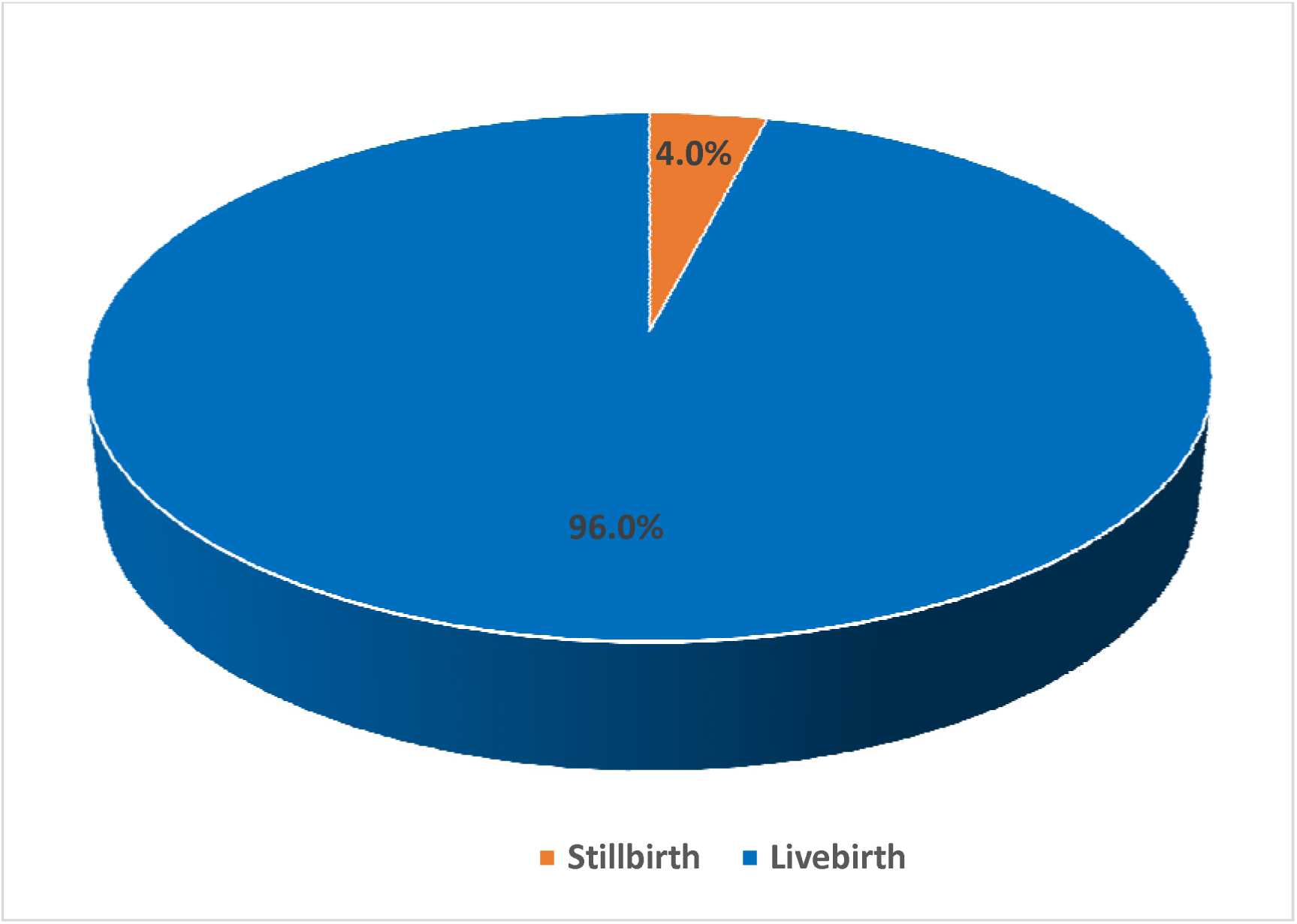
Prevalence of stillbirths in a semi-rural municipality of Ghana.

### Sociodemographic characteristics of study participants

The frequency distribution of sociodemographic characteristics of study participants are illustrated in Table 2. The average age of the mothers was 27.6 (sd=6.0) years, while nine percent of all the mothers were teenagers. A large proportion of the participants could not read or write (no formal education) (43%) and were not employed (64%). Furthermore, more than two-thirds of the mothers were from the Dagomba ethnic group (81%) and were married (93%). Finally, Islam was the most practised religion by the study participants (88%).

**Table 2:** Sociodemographic characteristics of study participants.

| Variables | Frequency (N) | Proportion (%) |
| --- | --- | --- |
| **Age category of mother** |  |  |
| < 20 years | 38 | 9.0 |
| 20 – 35years | 340 | 80.6 |
| > 35 years | 44 | 10.4 |
| Mean = 27.63 (SD=6.02) |  |  |
| **Marital status of mother** |  |  |
| Married | 391 | 92.7 |
| Not married | 31 | 7.3 |
| **Education level of mother** |  |  |
| No education | 182 | 43.1 |
| Primary school | 58 | 13.8 |
| High school | 138 | 32.7 |
| Tertiary | 44 | 10.4 |
| **Ethnicity of mother** |  |  |
| Dagomba | 340 | 80.6 |
| Gonja | 22 | 5.2 |
| Frafra/Grusi | 34 | 8.1 |
| Others (Asante, Ewe, Nigeria) | 26 | 6.1 |
| **Religion of mother** |  |  |
| Muslim | 372 | 88.2 |
| Christian | 50 | 11.8 |
| **Occupation of mother** |  |  |
| Unemployed | 269 | 63.7 |
| Employed | 153 | 36.3 |
| **Sex of baby (foetus)** |  |  |
| Male | 210 | 49.8 |
| Female | 212 | 50.2 |

### Obstetric and medical characteristics of study participants

The frequency distribution of medical and obstetric (antenatal) characteristics of respondents are shown in Table 3. Maternal anaemia was highest in the second (71%), followed by the first (64%) and third (45%) trimesters of pregnancy. Few of the mothers attended antenatal clinic at least eight times (28%), ingested sulphadoxine-pyrimethamine at least three times (36%), and did not use insecticide-treated bed nets (ITNs) during pregnancy (36%). Nearly three-quarters of the mothers were multiparous (71%) and multigravidae (74%). Additionally, slightly above a third of the mothers tested positive for sickle cell (38%), whereas almost 90% had positive blood rhesus status. A small proportion of the mothers had pre-eclampsia/eclampsia (2%), were diagnosed with antepartum haemorrhage (7%), were immunised against tetanus-diphtheria (7%), and had glucose-6-phosphate dehydrogenase (G6PD) defect (27%) during the period of pregnancy.

**Table 3:** Medical and obstetric characteristics of study participants.

| Variables | Frequency (N) | Proportion (%) |
| --- | --- | --- |
| **First-trimester body mass index (BMI)** |  |  |
| Underweight | 13 | 3.1 |
| Normal BMI | 279 | 66.1 |
| Overweight | 130 | 30.8 |
| **Number of antenatal care visits** |  |  |
| < 8 visits | 304 | 72.0 |
| ≥ 8 visits | 118 | 28.0 |
| **Sulphadoxine-pyrimethamine intake** |  |  |
| None | 25 | 5.9 |
| 1 – 3 doses | 247 | 58.6 |
| > 3 doses | 150 | 35.5 |
| **Pre-eclampsia/Eclampsia** |  |  |
| Yes | 9 | 2.1 |
| No | 413 | 97.9 |
| **Antepartum haemorrhage** |  |  |
| Yes | 34 | 8.1 |
| No | 388 | 91.9 |
| **Pre-pregnancy family planning use** |  |  |
| Yes | 75 | 17.8 |
| No | 347 | 82.2 |
| **Insecticide-treated bed nets use during pregnancy** |  |  |
| Yes | 270 | 64.0 |
| No | 152 | 36.0 |
| **Malaria episode during pregnancy** |  |  |
| Yes | 59 | 14.0 |
| No | 363 | 86.0 |
| **Gravidity** |  |  |
| Primigravidae | 112 | 26.5 |
| Multigravidae | 310 | 73.5 |
| **Parity** |  |  |
| Primiparity | 122 | 28.9 |
| Multiparity | 300 | 71.1 |
| **Mode of delivery** |  |  |
| Vaginal delivery | 379 | 89.8 |
| Caesarean section | 43 | 10.2 |
| **Gestational age at delivery** |  |  |
| < 37 weeks (preterm) | 91 | 21.6 |
| ≥ 37 weeks (normal term) | 331 | 78.4 |
| **First-trimester anaemia** |  |  |
| Anaemia | 268 | 63.5 |
| No anaemia | 154 | 36.5 |
| **Second-trimester anaemia** |  |  |
| Anaemia | 301 | 71.3 |
| No anaemia | 121 | 28.7 |
| **Third-trimester anaemia** |  |  |
| Anaemia | 191 | 45.2 |
| No anaemia | 231 | 54.8 |
| **Blood rhesus type** |  |  |
| Positive | 379 | 89.8 |
| Negative | 43 | 10.2 |
| **Sickle cell status** |  |  |
| Positive | 159 | 37.8 |
| Negative | 263 | 62.2 |
| **G6PD status** |  |  |
| Normal | 309 | 73.2 |
| Complete/partial defect | 113 | 26.8 |
| **TD immunization** |  |  |
| Immunized | 31 | 7.4 |
| Not immunized | 391 | 92.6 |

*G6PD: Glucose-6-phosphate dehydrogenase TD: Tetanus-diphtheria*

### Bivariate association of stillbirths with background variables

The bivariate association between stillbirths and the background variables are exhibited in Table 4. Among the sociodemographic, medical and obstetric or antenatal characteristics (variables) of the study participants, only sulphadoxine-pyrimethamine intake (p=0.012), insecticide-treated bed net use (p<0.001), parity (p=0.035), third-trimester anaemia status (p=0.030) and sickle cell status (p=0.003) were significantly associated with birth outcome prognosis (stillbirths) during bivariable analysis.

**Table 4:** Bivariate association of stillbirths with background variables.

| Variables | Bivariate analysis |  |  |
| --- | --- | --- | --- |
|  | Stillbirths (%) | Livebirths (%) | p-value <sup>a</sup> |
| ***Sociodemographic variables*** |  |  |  |
| **Age category of mother** |  |  |  |
| < 20 years | 1 (5.9) | 37 (9.2) | 0.573 |
| 20 – 35years | 13 (76.5) | 327 (80.7) |  |
| > 35 years | 3 (17.6) | 41 (10.1) |  |
| Mean = 27.63 (SD=6.02) |  |  |  |
| **Marital status of mother** |  |  |  |
| Married | 15 (88.2) | 376 (92.8) | 1.000 |
| Not married | 2 (11.8) | 29 (7.2) |  |
| **Education level of mother** |  |  |  |
| No education | 7 (41.2) | 175 (43.2) | 0.314 |
| Primary school | 1 (5.9) | 57 (14.1) |  |
| High school | 9 (52.9) | 129 (31.8) |  |
| Tertiary | 0 (0.0) | 44 (10.9) |  |
| **Ethnicity of mother** |  |  |  |
| Dagomba | 13 (76.5) | 327 (80.7) | 0.354 |
| Gonja | 0 (0.0) | 22 (5.4) |  |
| Frafra/Grusi | 3 (17.7) | 31 (7.7) |  |
| Others (Asante, Ewe, Nigeria) | 1 (5.9) | 25 (6.2) |  |
| **Religion of mother** |  |  |  |
| Muslim | 13 (76.5) | 359 (88.6) | 0.158 |
| Christian | 4 (23.5) | 46 (11.4) |  |
| **Occupation of mother** |  |  |  |
| Unemployed | 16 (94.1) | 253 (62.5) | 0.303 |
| Employed | 1 (5.9) | 152 (37.5) |  |
| **Sex of baby (foetus)** |  |  |  |
| Male | 6 (35.3) | 204 (50.4) | 0.322 |
| Female | 11 (64.7) | 201 (49.6) |  |
| ***Medical and obstetric variables*** |  |  |  |
| **First-trimester body mass index (BMI)** |  |  |  |
| Underweight | 1 (5.9) | 12 (3.2) | 0.423 |
| Normal BMI | 12 (64.7) | 267 (65.9) |  |
| Overweight | 5 (29.4) | 125 (30.9) |  |
| **Number of antenatal care visits** |  |  |  |
| < 8 visits | 14 (82.3) | 290 (71.6) | 0.333 |
| ≥ 8 visits | 3 (17.7) | 115 (28.4) |  |
| **Sulphadoxine-pyrimethamine intake** |  |  |  |
| None | 2 (11.8) | 23 (5.7) | **0.012<sup>\*</sup>** |
| 1 – 3 doses | 12 (70.6) | 235 (58.0) |  |
| > 3 doses | 3 (17.6) | 147 (36.3) |  |
| **Pre-eclampsia/Eclampsia** |  |  |  |
| Yes | 2 (11.8) | 7 (1.7) | 0.768 |
| No | 15 (88.2) | 398 (98.3) |  |
| **Antepartum haemorrhage** |  |  |  |
| Yes | 4 (23.5) | 30 (7.4) | 0.622 |
| No | 13 (76.5) | 375 (92.6) |  |
| **Pre-pregnancy family planning use** |  |  |  |
| Yes | 3 (17.7) | 72 (17.8) | 0.989 |
| No | 14 (82.3) | 333 (82.2) |  |
| **Insecticide-treated bed nets use during pregnancy** |  |  |  |
| Yes | 4 (23.5) | 266 (65.7) | < 0.001* |
| No | 13 (76.5) | 139 (34.3) |  |
| **Malaria episode during pregnancy** |  |  |  |
| Yes | 1 (5.9) | 58 (14.3) | 0.326 |
| No | 16 (94.1) | 347 (85.7) |  |
| **Gravidity** |  |  |  |
| Primigravidae | 2 (11.8) | 110 (27.2) | 0.051 |
| Multigravidae | 15 (88.2) | 295 (72.8) |  |
| **Parity** |  |  |  |
| Primiparity | 3 (17.7) | 119 (29.4) | 0.035* |
| Multiparity | 14 (82.3) | 286 (70.6) |  |
| **Mode of delivery** |  |  |  |
| Vaginal delivery | 15 (88.2) | 364 (89.9) | 0.827 |
| Caesarean section | 2 (11.8) | 41 (10.1) |  |
| **Gestational age at delivery** |  |  |  |
| < 37 weeks (preterm) | 4 (23.5) | 87 (21.5) | 0.822 |
| ≥ 37 weeks (normal term) | 13 (76.5) | 318 (78.5) |  |
| **First-trimester anaemia** |  |  |  |
| Anaemia | 11 (64.7) | 257 (63.4) | 0.554 |
| No anaemia | 6 (35.3) | 148 (36.6) |  |
| **Second-trimester anaemia** |  |  |  |
| Anaemia | 13 (76.5) | 288 (71.1) | 0.700 |
| No anaemia | 4 (23.5) | 117 (28.9) |  |
| **Third-trimester anaemia** |  |  |  |
| Anaemia | 13 (76.5) | 178 (44.0) | 0.030* |
| No anaemia | 4 (23.5) | 227 (55.0) |  |
| **Blood rhesus type** |  |  |  |
| Positive | 15 (88.2) | 364 (89.9) | 0.455 |
| Negative | 2 (11.8) | 41 (10.1) |  |
| **Sickle cell status** |  |  |  |
| Positive | 7 (41.2) | 152 (37.5) | 0.003* |
| Negative | 10 (58.8) | 253 (62.5) |  |
| **G6PD status** |  |  |  |
| Normal | 14 (82.4) | 295 (72.8) | 0.732 |
| Complete/partial defect | 3 (17.6) | 110 (27.2) |  |
| **TD immunization** |  |  |  |
| Immunized | 3 (17.7) | 28 (6.9) | 0.121 |
| Not immunized | 14 (82.3) | 377 (93.1) |  |

\* $p < 0.05$
<sup>a</sup> Chi-square/Fisher's exact test G6PD: Glucose-6-phosphate dehydrogenase TD: Tetanus-diphtheria

### Maternal factors associated with stillbirths

The maternal factors associated with stillbirths are presented in Table 5. In the multiple logistic regression analysis, three variables particularly, insecticide-treated bed net use, third-trimester anaemia status and sickle cell status, remained statistically significant in the final model. The model revealed that the odds of mothers with anaemia in the third trimester of pregnancy having stillbirths was 3.15 times (95%CI: 2.94 – 6.10; p=0.034) compared to the non-anaemic mothers. Moreover, mothers with positive sickle cell status had 2.91 (95%CI: 2.50 – 5.01; p=0.018) increased odds of stillbirths compared to their counterparts with negative sickle cell status. Finally, participants who did not sleep under insecticide-treated bed nets throughout pregnancy had 7.75 (95%CI: 4.33 – 8.80; p=0.001) higher odds of stillbirths than those who used it.

**Table 5:** Multiple logistic regression of maternal factors of stillbirths.

| Variables | Logistic regression analyses |  |  |  |
| --- | --- | --- | --- | --- |
|  | Univariate analysis |  | Multivariate analysis |  |
|  | cOR (95%CI) | p-value | aOR (95%CI) | p-value |
| **Sulphadoxine-pyrimethamine intake** |  |  |  |  |
| 1 – 3 doses |  | 1.000 |  | 1.000 |
| > 3 doses | 0.11 (0.15 – 0.88) | 0.037* | 0.17 (0.02 – 1.35) | 0.094 |
| None | 1.46 (0.31 – 6.82) | 0.631 | 2.68 (0.41 – 7.13) | 0.301 |
| **Insecticide-treated bed nets use** |  |  |  |  |
| Yes |  | 1.000 |  | 1.000 |
| No | 6.22 (3.99 – 8.40) | 0.002* | **7.75 (4.33 – 8.80)** | **0.001\*** |
| **Parity** |  |  |  |  |
| Primiparity |  | 1.000 |  | 1.000 |
| Multiparity | 6.66 (3.87 – 7.70) | 0.047* | 4.64 (0.57 – 7.60) | 0.151 |
| **Third-trimester anaemia** |  |  |  |  |
| No anaemia |  | 1.000 |  | 1.000 |
| Anaemia | 4.02 (2.29 – 5.12) | 0.017* | **3.15 (2.94 – 6.10)** | **0.034\*** |
| **Sickle cell status** |  |  |  |  |
| Negative |  | 1.000 |  | 1.000 |
| Positive | 1.51 (1.10 – 6.41) | 0.019* | **2.91 (2.50 – 5.01)** | **0.018\*** |
| **Regression model** |  |  |  |  |
| $R^2$ | 0.6030 | | | |
| $p$ value | < 0.001 | | | |
| * $p < 0.05$ | cOR: crude odds ratio | aOR: adjusted odds ratio | Bold: significant aOR, p-value, and 95%CI for risk factors | |

## Discussion

The study assessed the prevalence and contributing factors of stillbirths in a semi-rural locality. The prevalence of stillbirths in the present study was found to be 4.0%, which is in line with a multicenter study in Nigeria (11). This similarity might be due to the uniformity in the healthcare system and provision of services for pregnant mothers in Ghana and Nigeria. Our study’s stillbirth rate was also lower than the estimation in a retrospective cohort study conducted in a tertiary facility in Southern Ghana (10.6%) (20) but higher than the study findings from China (0.9%) and South Africa (3.5%) (10,17). The discrepancy might be due to variations in the study area and the inclusion and exclusion criteria (methodologies) used in the previous studies. In addition, several unreported cases of stillbirths in the community and/or municipal health facilities could be responsible for this variation. In comparison, China and South Africa have advanced and modernised healthcare systems that offer well-tailored services to pregnant women, thereby contributing to the reduction of stillbirth rates in these countries. Therefore, it could indicate that the care for women during antenatal and labour (intrapartum) might not have received appropriate and standardized obstetric practices and protocols, leading to higher stillbirth incidence in this semi-rural Ghana. On the other side, the lesser prevalence of stillbirths in this study, compared to Southern Ghana, might be due to the deployment of trained traditional birth attendants as agents of facility deliveries and trained community health nurses to perform the duties of midwives in the Savelugu municipality of Northern Ghana. This initiative by the Catholic Relief Services (development partner) was adopted by the Northern Regional Health Directorate of Ghana Health Service to improve maternal and perinatal health outcomes, which might have contributed to the reduction of the stillbirth rate in Northern Ghana. Nonetheless, the relatively elevated incidence of stillbirths in the study area is still alarming as postpartum mothers and their families face psychological trauma after experiencing stillbirths (11,37) during or following a nine-month journey of pregnancy.

In this study, maternal anaemia in the third trimester of pregnancy was independently associated with stillbirths. Whilst a retrospective study in Columbia (38) was in agreement with the present study, a similar study in Pakistan (39) was not in support. Anaemia is known to be a significant predictor of malnutrition among pregnant mothers as iron deficiency type is apparently the most prevalent cause of anaemia during gestation (40). As anaemia causes inadequate oxygen and nutrient supply to the foetus during the late stage of pregnancy (41,42), it could lead to a deficient supply of these nutrients and oxygen to the foetus which subsequently causes hypoxic death of the foetus either in-utero, during childbirth or within a short period after birth (43). To prevent gestational anaemia and subsequent stillbirths, it is critical to strengthen public health initiatives and clinical protocols (15). These actions could be achieved through early screening and diagnosis, prompt patient referral, integrated nutrition counselling, routine iron-folate supplementation and culturally tailored community-based education promoting iron-folate concordance and dietary quality (44).

This study also found that mothers with positive sickle cell status had increased odds of stillbirth delivery. The observed findings align with previous research indicating severe pregnancy and childbirth complications for both mothers and newborns affected by sickle cell (45). These complications may stem from the physiological adaptations inherent in the genetic makeup of pregnant women with sickle cell trait or disease (45,46). Such adaptations can impact the cardiovascular, haematologic, respiratory, and renal structures in the materno- foetal system, which can impair the overall health and ability to sustain a healthy pregnancy (46). Consequently, these physiological challenges may contribute to the reduced likelihood of delivering a healthy newborn. It therefore becomes imperative for healthcare professionals especially midwives and medical doctors to recognise this risk and adopt appropriate measures by providing targeted and special care for pregnant women affected by sickle cell.

Mothers who did not sleep under insecticide-treated bed nets during pregnancy were more likely to deliver stillbirths. In line with the present findings, mothers’ sleep practices, particularly those who did not sleep under insecticide-treated bed nets during pregnancy, were prone to stillbirths in a cross-sectional study in Ghana (12) but were contrasting to a meta-analysis by Flenady and his colleagues (47). Despite the constant supply of free insecticide-treated bed nets to pregnant mothers for malaria prevention during their first ANC visits in Ghana (28), it is reported that most mothers do not sleep under the nets due to unfavourable (hot) weather conditions in Northern Ghana, while others also use the nets for subsistence farming purposes in their homes (48). The non-use of the bed nets could expose them to malaria infections and subsequent anaemia in pregnancy, thereby increasing their risk of stillbirth delivery (49). This underscores the need for expanded and sustained health education campaigns including outreach activities in both health facilities and community settings to foster insecticide-treated bed nets utilisation in semi-rural areas of Ghana.

Multiparity was found to be associated with stillbirths in the semi-rural community during the regression analysis. Evidence from Ethiopia-based comparative studies (50,51) corroborates the results of the current investigation. Similarly, in a cross-sectional study from Nigeria, multiparity was associated with stillbirths (11). Most multiparous women perceive themselves as experienced and often delay or underutilise ANC services during pregnancy, which may reduce opportunities for early detection of complications associated with stillbirths. Furthermore, multiparity with short interpregnancy intervals results in nutritional deficiencies such as iron, folate and vitamin B12, which could lead to foetal growth restrictions and stillbirths (1,52). While promoting family planning initiatives across communities remains essential in mitigating multiparity (53,54), prioritising diverse targeted interventions for multiparous women is equally critical in efforts to prevent the risk of stillbirths in the municipality.

Interestingly, a large proportion of the study participants who had positive sickle cell status and did not use insecticide-treated bed nets also made less than eight ANC visits and had anaemia during each trimester of pregnancy. This implies that fewer ANC visits and low haemoglobin levels in each trimester during pregnancy could precipitate suboptimal intra- uterine and gestational environments (41,42) leading to an increased maternal risk for stillbirth delivery (49,55) in the municipality. Therefore, greater attention should be paid to anaemia in pregnancy and ANC visits to strengthen maternal and child health outcomes in the country.

### Strengths and limitations of the study

The study recorded some limitations. First, the prevalence of stillbirths in the municipality may be underestimated or overestimated since the study participants were sampled from six-month cross-sectional retrospective data from health facilities. There could be potential misreporting (underreporting), misrecording and misclassification of stillbirths and other clinical (health) data like sickle cell determination by healthcare professionals (27,56,57), which could affect the estimates of stillbirths and positive sickle cell in the municipality. The data also lacked details on whether the stillbirths happened in the antepartum or intrapartum stages of delivery. The absence of this information could imply a considerable blend of antepartum and intrapartum stillbirths in the dataset. Additionally, the use of retrospective data (such as sickle cell, maternal weight, multiparity, haemoglobin levels, and gestational age) from records review (MCHRBs and delivery registers) could be a limitation of the study as a result of possible classification, measurement and documentation errors (56). It is difficult to ascertain whether the necessary standard protocols were adequately adhered to before measurement, potentially affecting the dataset’s quality in the analysis. Notwithstanding, the stillbirth estimates and measurements represent the available information in the municipal health facilities which are used for decision-making in the municipality.

The study also faced limitations in generalising the maternal factors of stillbirths because it relied on cross-sectional data. Therefore, the findings of this study could not be used to draw causal inferences but rather to highlight significant associations of stillbirths to the scientific community. Furthermore, the reliability of self-reported data may be affected by recall bias among study participants, posing a methodological limitation in the study. Ultimately, the study lacked information on certain variables related to the independent (exposure) variables. Specifically, maternal data on smoking status, alcohol intake and socioeconomic status were not collected. Despite this, the study provided significant findings for potential interventions in the municipality and the broader semi-rural Ghana.

On the other hand, the study encountered some notable strengths. At the outset, the study addresses a significant gap in the maternal and perinatal health literature by quantitatively examining the association between maternal factors and stillbirths. This directly connects maternal health factors to perinatal health. The study goes beyond reporting univariate and bivariate associations by also conducting multivariable analysis to identify maternal factors of stillbirths, resulting in a more rigorous and robust set of findings.

## Conclusion

The prevalence of stillbirths is relatively high in the Savelugu municipality. These stillbirths are independently influenced by medical factors, particularly anaemia and positive sickle cell statuses and unsafe health practices such as non-use of ITNs during pregnancy. Timely identification of these factors among pregnant women (anaemia and sickle cell status) is vital to prompt early intervention strategies in Ghanaian health facilities. This could be achieved by strengthening obstetric triage at ANC clinics and labour/delivery rooms and also encouraging early referrals to higher levels of care. Secondly, due to the frequent misclassification of most perinatal and neonatal deaths as stillbirths (21), thorough perinatal death audits, which aim to identify the cause and contributing factors of stillbirths, should be promoted in healthcare facilities in semi-rural Ghana.

Additionally, the management of Ghana Health Service (especially municipal/district health directorates), alongside program planners and other stakeholders involved in enhancing perinatal and maternal health, should also be aware of these maternal factors contributing to stillbirths. These health authorities should launch and implement effective programs, including robust referral systems through training and capacity building of health care providers and community cadres to facilitate early referral and management. Policy efforts should further be targeted at promoting enhanced health education sessions on undernutrition (anaemia) and the regular use of ITNs during pregnancy at facility ANC clinics and communities (via community durbars) in semi-rural Ghana. Finally, we recommend future research to incorporate other maternal variables, such as socioeconomic status, prenatal smoking and alcohol intake, to thoroughly examine and provide a more comprehensive understanding of the maternal factors contributing to stillbirths in both the municipality and the wider Northern Region.

## List of abbreviations

ANC: Antenatal care
BMI: Body mass index
G6PD: Glucose-6-phosphate dehydrogenase
ITNs: Insecticide-treated bed nets
MCHRBs: Maternal and child health record books
STATA: Statistics and data
TD: Tetanus-diphtheria
VIF: Variance inflation factor
WHO: World Health Organization

## Declarations

### Ethics approval and consent to participate

This study was conducted according to the guidelines laid down in the Declaration of Helsinki, and all procedures involving research study participants were approved by the Navrongo Health Research Centre Institutional Review Board with approval number NHRCIRB373. Permission was also granted by the Ghana Health Service at the municipal directorate and health facilities level. Written informed consent/assent was obtained from all study participants and/or legal representatives and also from parents/guardians of participants under 18 years of age. All participants were able to understand, read and write about consent to participate.

### Consent for publication

Not applicable

## Data Availability

The datasets collected, generated, and/or analyzed during the present study are available from the corresponding author upon reasonable request. Data sharing not applicable to this article as no datasets were generated or analyzed during the current study.

## Acknowledgements

The authors are thankful to all research assistants, study participants, and volunteers who contributed in diverse ways to the successful implementation of the study. We are also grateful to Elvis Brown Ayaala, Abdulai Sulemana, Allor Mohammed, and Julien Beweleyir for their supportive supervision and data collection exercise during this study.

## Funding statement

There was no funding support

## Conflict of interest statement

The authors declared that they have no conflict of interest.

## Availability of data and materials

The datasets collected, generated, and/or analysed during the present study are available from the corresponding author upon reasonable request. Data sharing not applicable to this article as no datasets were generated or analysed during the current study.

## Authors’ contributions

SAG, AA, MG, CSD and PAA designed and conceptualised the study. SAG, AA, MG, WP, CA and CSD carried out the data collection while SAG, WP, CA and PAA analysed and interpreted the data. All authors contributed to drafting the initial manuscript, reviewing and approving the final manuscript.

## Authors’ acronyms

Silas Adjei-Gyamfi (SAG); Abigail Asirifi (AA); Mamudu Gafaru (MG); Wisdom Peprah (WP); Clotilda Asobuno (CA); Constance Siakwan Dapaah (CSD); Paul Armah Aryee (PAA)

## Notes

### Competing Interest Statement

The authors have declared no competing interest.

### Author Declarations

This study was conducted according to the guidelines laid down in the Declaration of Helsinki, and all procedures involving research study participants were approved by the Navrongo Health Research Centre Institutional Review Board with approval number NHRCIRB373. Permission was also granted by the Ghana Health Service at the municipal and health facilities level. Written informed consent/assent was obtained from all study participants and/or legal representatives and also from parents/guardians of participants under 18 years of age.

### Summary of Updates

Some revisions have been made throughout the entire manuscript.

